# Implications of red state/blue state differences in COVID-19 death rates

**DOI:** 10.1101/2022.04.08.22273628

**Authors:** Mary L. Adams

**Affiliations:** On Target Health Data LLC, Suffield CT

## Abstract

The study objective was to explore state death rates pre- and post- 4/19/2021 (date vaccines were assumed available) and the relative contributions of 3 factors to state death rates post- 4/19/2021: 1) vaccination rates, 2) prevalence of obesity, hypertension, diabetes, COPD, cardiovascular disease, and asthma and 3) red vs. blue states, to better understand options for reducing deaths. The ratio of red to blue state deaths/million was 1.6 pre-4/19/2021 and 2.3 between 4/19 and 2/28/2022 resulting in >222,000 extra deaths in red states or 305/ day. Adjusted betas from linear regression showed state vaccination rates had the strongest effect on death rates while red vs. blue states explained more of the difference in state death rates (60% vs. 46% for vaccination rates) with mean vaccination rates ~10% higher in blue states. Results suggest that increasing vaccination rates in red states could potentially save thousands of lives as the pandemic continues.

## Background

The cost of COVID-19 was estimated at $16 trillion (*1*) through the fall of 2021 and the death toll to 2/28/2022 was 943,775 and climbing, even with effective vaccines available (*2*). Obesity, hypertension, diabetes, COPD, cardiovascular disease, and asthma can affect death rates (*3, 4*) and higher death rates have been noted in red vs. blue states (*5*). The study objective was first to explore state death rates pre- and post- 4/19/2021 and second to use linear regression to explore the contributions of 3 factors to state death rates post- 4/19/2021: 1) vaccination rates, 2) prevalence of the six chronic conditions and 3) red vs. blue states, to better understand opportunities for reducing deaths.

## Methods

COVID-19 deaths and the percentage of fully vaccinated adults on selected dates for each state were obtained from USA Facts (*2*). The date of 3/1/2020 was chosen as the beginning of the pandemic,4/19/2021 was chosen when vaccines were available, and adult vaccination rates were as of 8/27/2021. Publicly available 2019 Behavioral Risk Factor Surveillance System (BRFSS) data (*6*) from 409,810 adults from 49 states (no NJ) and DC, were used to obtain state prevalence rates for obesity, hypertension, diabetes, COPD, cardiovascular disease, and asthma (*3,4*). Data were weighted to be representative of the total adult population of each state. Median survey response rate was 49.4% (*7*). States were coded as red (Trump) or blue (Clinton) based on voting in the 2016 election.

Number of deaths per day and mean deaths/million population pre- and post- 4/19 were computed separately for red and blue states in Excel and compared. Stata version 14.2 was used for other analysis, using the survey option to account for the complex survey design of the BRFSS unless otherwise noted. Regression models included the 3 factors together and each factor separately to estimate coefficients and R^2^. Regression was done with the survey option and repeated without the survey option to obtain standardized betas.

## Results

Total deaths (not per million) indicated that daily deaths decreased in blue states from 588/day pre-4/19/2021 to 373/day from 4/19-2/28/2022 while the number of daily deaths in red states increased from 769/day to 840/day in the same time periods. The change in blue states represents a 36% reduction in deaths/day and the change in red states represents a 9% increase resulting in >222,000 excess deaths in red states over 729 days or 305/day. Taking the different state populations into account, the ratio of red to blue state deaths/million was 1.6 pre-4/19/2021 and 2.3 between 4/19 and 2/28/2022. Highest mean death rates post 4/19 were for states with <55% of adults fully vaccinated (1,572 deaths/million) and for red states (1,403 deaths/million) with lowest rates for blue states (824 deaths/million) and states with >66.6% adults fully vaccinated (874 deaths/million) with all values for intermediate vaccination rates and different number of risks between 1,086 and 1,275 deaths/million. Factors tended to be associated, with no blue states having <55% adults fully vaccinated and no red states having >66.6% fully vaccinated, with mean percentages of the <u>un</u>vaccinated at 31.5% in blue states and 42.4% in red states. Less dramatic associations were found between vaccination and risk, and risk and red/blue states. Unadjusted results comparing red and blue states on a wide range of factors are shown in Table 1; adults in red states had significantly higher prevalence rates for all measures listed except asthma and higher vaccination rates. Summary results for various regression models (Table 2) show vaccination rates, with standardized beta of 0.52, had the strongest effect on death rates while red vs. blue states explained more of the difference in state death rates (60% vs. 46% for vaccination rates). R^2^ values for all models except just the COVID risks ranged from 0.46 for just vaccination rate to 0.62 for the model with all 3 factors and not requesting adjusted betas. Changes in selected dates did not substantially alter results.

**Table 1.** Comparing red vs. blue states on selected measures potentially related to COVID deaths; 2019 Behavioral Risk Factor Surveillance System, except vaccine data*

| Measure | Blue states<sup>†</sup> |  | Red states<sup>†</sup> |  |
| --- | --- | --- | --- | --- |
|  | % | 95% CI | % | 95% CI |
| Composite COVID risk measure |  |  |  |  |
| Obesity | 28.1 | 27.7-28.6 | 33.6 | 33.2-34.1 |
| Diabetes | 10.0 | 9.8-10.3 | 11.8 | 11.5-12.0 |
| Hypertension | 29.9 | 29.5-30.3 | 34.5 | 34.1-34.9 |
| COPD<sup>§</sup> | 5.3 | 5.1-5.5 | 7.5 | 7.4-7.7 |
| Heart disease | 5.4 | 5.2-5.6 | 6.9 | 6.7-7.1 |
| Current asthma | 8.9 | 8.7-9.2 | 9.0 | 8.8-9.2 |
| Behavioral risk factors associated with above |  |  |  |  |
| Current smoking | 12.5 | 12.2-12.8 | 17.3 | 17.0-17.7 |
| Sedentary | 23.8 | 23.4-24.2 | 27.6 | 27.3-28.0 |
| Not meet activity recommendations | 50.6 | 50.1-51.1 | 55.5 | 55.1-56.0 |
| Eat <5 a day | 84.4 | 84.1-84.8 | 86.8 | 86.5-87.1 |
| All overweight | 63.7 | 63.2-64.2 | 68.6 | 68.2-69.0 |
| Other factors potentially related to COVID deaths |  |  |  |  |
| Any disability | 24.9 | 24.5-25.3 | 30.1 | 29.8-30.5 |
| Fair/poor health | 17.0 | 16.7-17.3 | 19.8 | 19.5-20.1 |
| Uninsured | 11.1 | 10.8-11.4 | 14.4 | 14.0-14.7 |
| Cost barrier to care | 11.7 | 11.4-12.0 | 14.7 | 14.4-15.0 |
| No regular MD | 22.2 | 21.8-22.6 | 24.2 | 23.8-24.6 |
| No flu shot past year | 55.9 | 55.4-56.3 | 58.0 | 57.6-58.4 |
| <55% fully vaccinated | 0 |  | 28.3 | 28.2-28.5 |
| >66.6% fully vaccinated | 59.6 | 59.2-60.1 | 0 |  |

\* USA Facts as of 8/27/2021.
<sup>†</sup> Red and blue states based on voting in 2016 Presidential election
<sup>§</sup> Chronic obstructive pulmonary disease

**Table 2.** Summary of regression results from Stata for combined 2019 B and USA Facts data; dependent variable= US COVID deaths 4/19/2021-2/28/2022. N=49 states+ DC (no NJ).

| Variable(s) | Coefficient | R<sup>2</sup> | Adjusted beta |
| --- | --- | --- | --- |
| Just % unvaccinated** | 34.78 | 0.46 | n.a. |
| Just red vs blue states† | 578.69 | 0.60 | n.a. |
| Just COVID risks§ | 27.43 | 0.01 | n.a. |
| All 3 factors without survey option |  |  |  |
| % Unvaccinated** | 23.78 | 0.53 | 0.52 |
| Red vs. blue states† | 179.77 | 0.53 | 0.24 |
| COVID risks § | 12.52 | 0.53 | 0.04 |
| All 3 using survey option |  |  |  |
| % Unvaccinated** | 11.29 | 0.62 | n.a. |
| Red vs. blue states† | 454.19 | 0.62 | n.a. |
| COVID risks § | 7.89 | 0.62 | n.a. |

\* Behavioral Risk Factor Surveillance System
\*\* As of 8/27/2021 USA Facts
† Red and blue states based on voting in 2016 Presidential election
§ Obesity, diabetes, hypertension, chronic obstructive pulmonary disease, heart disease, asthma

## Discussion

Once vaccines were available, they had the strongest effect on death rates among the 3 factors, which is consistent with results that found vaccine effectiveness against death at 99% (98.5-99.6%) for fully vaccinated individuals (*8*). As much as 62% of the difference between state COVID death rates was explained by the 3 factors combined, with 46% explained by just vaccination rates. Second in strength of effect on death rates was the variable representing red or blue states, which explained 60% of the difference in state death rates but represents a “black box” of factors. While these results don’t identify all the differences between red and blue states, the measures in Table 1 offer some key differences: vaccination rates (which by themselves explained 46% of death rate differences), chronic conditions associated with higher COVID death rates, access to health care, general health status, disability, and receipt of a recent influenza vaccination were all worse in red states. Other possible factors affecting the death rates that likely differed between red and blue states include ways the pandemic was handled, including mask mandates, lockdowns, and other restrictions. Taken together, these differences resulted in a death rate/million in red states post 4/19/2021 that was 2.3 times that in blue states, an extra 222,000 deaths overall, and a 9% increase in death rates compared with the pre-vaccine period, at a time when the death rate in blue states decreased 36%.

There are several limitations to this study. Data on COVID deaths include deaths in nursing homes while the BRFSS data only include non-institutionalized adults. The impact of this limitation can’t be measured but is likely to be greater in the period prior to April 19 when the pandemic hit nursing homes hard. The data for vaccinations do not distinguish booster shots or any differences in the effectiveness of different brands. It is also difficult to determine (beyond the standardized beta results) which differences between red and blue states have the most influence on death rates or how those factors might be modified to lower death rates.

Perhaps putting these results in context with other studies might help. A study of 30 industrialized countries (*9*) found that obesity, population density, the age structure of the population, population health, GDP, ethnic diversity, and how the pandemic was handled explained 63% of the intercountry variation in COVID death rates. That regression R^2^ was very similar to the R^2^ of 0.60 for just red vs. blue states and the R^2^ of 0.62 for the study model with all 3 factors but the global study excluded vaccination rates. Population density, age and racial composition, pandemic response, etc. are likely among the differences between red and blue states that contribute to that coefficient of determination of 0.60. From this list, only the handling of the pandemic seems amenable to modification, along with vaccination rates noted from this study. Given possible alternatives, increasing vaccination rates especially in red states seems the best option to reduce COVID deaths. Vaccine promotions may need to be carefully tailored to motivate this group. Success may be limited because differences will remain between red and blue states that can affect death rates, as they likely affected the differences in death rates seen before 4/19/2021.

Over the course of the pandemic through 2/28/2022, excess deaths in red states resulted in > 220,000 more deaths compared with blue states, or 305 deaths/day. Deaths are not the only cost of a pandemic (*1*), but this study did not compare dollar or other costs between red and blue states. Other aspects of the pandemic such as the impact on healthcare systems and the economy may also be quite different for red and blue states.

## Data Availability

BRFSS Data available at: https://www.cdc.gov/brfss/annual_data/annual_2019.html.

Death Data: were available at https://usafacts.org/ but can be obtained from author

https://www.cdc.gov/brfss/annual_data/annual_2019.html.

